# Routine CBC-Derived Markers of Systemic Inflammation Are Associated With Growth of the Dilated Ascending Aorta During CT Surveillance

**DOI:** 10.64898/2026.11.05.26367393

**Authors:** Bruno Murad, Carlos Alberto Campello Jorge, Grace George, Ivo Queiroz Costa Neto, Yogesh Karnam, Gregory Spahlinger, Marion Hofmann Bowman, Nicholas S. Burris

## Abstract

**Background:** Ascending aortic surveillance relies on diameter thresholds, yet most dissections occur below them. Inflammation drives aortic wall remodeling independent of diameter. Inflammation drives aortic wall remodeling independent of diameter. Inflammatory indices derived from routine blood counts, such as neutrophil-to-lymphocyte ratio (NLR), red cell distribution width (RDW), and systemic immune-inflammation index (SII) have been show to predict adverse cardiovascular outcomes, but their association with ascending aortic growth is unclear.

**Methods and Results:** We retrospectively studied adults with ≥2 computed tomography (CT) scans of the native ascending aorta ≥6 months apart at a single center (2000–2023), excluding those with aortitis or no complete blood count during follow-up. Markers were summarized as 95th percentile values per patient and per scan interval. Growth was defined as a ≥2-mm increase in mid-ascending aortic diameter. Patient-level logistic regression and interval-level generalized estimating equation models were adjusted for age, sex, body mass index, baseline diameter, systolic blood pressure, diabetes, and heritable thoracic aortic disease. Among 3,262 patients, data were available in 1,882 for RDW, 1,471 for NLR, and 1,468 for SII; 192 (10.2%) met criteria for growth. Only RDW was independently associated with growth in the overall cohort (patient level: odds ratio [OR] 1.071, 95% CI 1.002–1.144, p=0.043; interval level: OR 1.075, 95% CI 1.012–1.141, p=0.019); NLR (p=0.080) and SII (p=0.164) were not. Among patients with guideline-defined ascending aortic dilation (≥40 mm), all three markers were associated with interval growth (RDW OR 1.14, NLR OR 1.05, SII OR 1.01 per 100 units; all p≤0.014), whereas none were among aortas <40 mm; effect modification was significant for NLR (p=0.01).

**Conclusions:** RDW was modestly associated with ascending aortic growth overall, and all three markers were associated with interval growth in patients with guideline-defined aortic dilation, suggesting that systemic inflammation may contribute to progression of aortic dilation. Prospective studies with more specific inflammatory markers are needed.

**Clinical Perspective:** *What Is New?:*

- Among 1,882 patients undergoing serial computed tomography surveillance, higher red cell distribution width was modestly but independently associated with ascending aortic growth, whereas the neutrophil-to-lymphocyte ratio and systemic immune-inflammation index were not associated with growth in the overall cohort.
- In patients with guideline-defined ascending aortic dilation (≥40 mm), all three inflammatory markers were associated with growth across surveillance intervals, a signal not seen in nondilated aortas.

*What Are the Clinical Implications?:*

- Routine blood count markers of inflammation may carry information about disease activity in patients under surveillance for ascending aortic dilation, although effect sizes are too modest for standalone risk stratification.
- Prospective studies with standardized inflammatory panels obtained at fixed surveillance intervals are needed to test whether inflammation identifies patients with dilated aortas who are at risk of growth.

## Introduction

Ascending thoracic aortic aneurysms (aTAA) are managed primarily through serial imaging surveillance, in which absolute aortic diameter is measured to predict the risk of life-threatening complications. However, more than 90% of dissections occur in patients below these surgical thresholds, with a median pre-dissection diameter of approximately 40 mm.^1–4^ This well-described paradox exposes a fundamental gap in current size-based risk stratification and underscores the need for complementary markers of disease activity that extend beyond aortic diameter.

Aortic aneurysm formation and progression are driven at the cellular level by degeneration of the medial layer of the aortic wall.^1^ Under pathological conditions, contractile vascular smooth muscle cells dedifferentiate into a synthetic phenotype, producing matrix metalloproteinases and proinflammatory factors that remodel the extracellular matrix and reduce structural integrity.^5,6^ This phenotypic switch can be induced by inflammatory cell infiltration and oxidative stress, and is accompanied by vascular inflammation independent of the degree of aortic dilation.^7^ Because these processes precede and may drive measurable diameter change, circulating markers that reflect systemic inflammatory burden have been proposed as potential surrogates of active aortic remodeling.

Several inexpensive, widely available indices derived from the complete blood count (CBC) have emerged as markers of systemic inflammation. The neutrophil-to-lymphocyte ratio (NLR) and the systemic immune-inflammation index (SII) have been linked to adverse cardiovascular outcomes and stroke risk.^8,9^ Red cell distribution width (RDW) has similarly emerged as a robust prognostic marker in cardiovascular disease, where elevated values are thought to reflect chronic inflammation, oxidative stress, and cytokine-mediated impairment of erythropoiesis.^10–12^ Despite the biological plausibility of a link between systemic inflammation and aortic remodeling, the value of these routinely collected markers for predicting growth of the ascending aorta during surveillance remains unclear.

In this study, we evaluated whether CBC-derived inflammatory markers are associated with interval growth of the ascending aorta among a large cohort of patients undergoing serial CT surveillance. We specifically aimed to determine whether these markers independently predict aortic growth at both the patient level and the individual surveillance-interval level, after adjustment for established demographic and clinical risk factors. We further examined whether these associations differed in patients with guideline-defined ascending aortic dilation, the population in whom imaging surveillance is recommended.

## Methods

The data that support the findings of this study are available from the corresponding author on reasonable request.

### Study Design and Population

This single-center, retrospective cohort study was conducted using data collected between January 2000 and December 2023 at the University of Michigan. The study received approval from the local Institutional Review Board (HUM00133798), which waived the requirement for informed consent. Reporting was performed in accordance with the Strengthening the Reporting of Observational Studies in Epidemiology (STROBE) guidelines.

Study participants were identified via institutional aortic research databases and electronic medical record (EMR) queries. Inclusion criteria comprised adult patients (≥18 years) with a minimum of two serial computed tomography (CT) scans of the ascending aorta, separated by at least six months, without prior surgical repair of the ascending aorta. Given that the average pre-dissection diameter is approximately 40 mm and risk is present below conventional aortic diameter boundaries,^2^ no minimum dilation threshold was imposed. Patients were excluded if they had a known diagnosis of aortitis or if no complete blood count (CBC) data were available during the surveillance period. Heritable thoracic aortic disease (HTAD) was defined as the presence of any of the following: Marfan syndrome, Loeys-Dietz syndrome, vascular Ehlers-Danlos syndrome or non-syndromic pathogenic variants (e.g., *ACTA2*, *MYH11*, or *MYLK*)

### Data Collection and Laboratory Integration

We extracted mid-ascending aortic (mid-AsAo) diameters from structured clinical radiology reports using an in-house text processing pipeline that has been employed in other related studies.^1^^3^ Of note, all aortic measurements were performed in a specialized 3D imaging lab, by trained image analysis technologists using standardized protocols with expert radiologist oversight. Implausible values and extreme interval changes suggestive of typographical errors were manually audited and corrected.

EMR data were abstracted for baseline demographics, comorbidities, vital signs, and clinical outcomes. Three CBC-derived inflammatory markers were evaluated: red cell distribution width (RDW), neutrophil-to-lymphocyte ratio (NLR), and systemic immune-inflammation index (SII). NLR was calculated as the absolute neutrophil count divided by the absolute lymphocyte count. SII was calculated as (platelet count × absolute neutrophil count) / absolute lymphocyte count. All absolute count values were normalized to K/µL prior to calculation. For SII and NLR computation, all components were required on the same calendar date.

Inflammatory marker values were summarized at two levels. At the patient level, each marker was summarized using the 95th percentile of all eligible CBC results obtained between the patient’s first and last CT scans, resulting in one marker value per patient across the entire surveillance period. At the interval level, each marker was summarized using the 95th percentile of all eligible CBC results obtained between each pair of consecutive CT scans, resulting in one marker value per imaging interval. Because the number of CBC draws varied across patients, each marker’s 95th percentile was computed by averaging the two nearest results, using midpoint interpolation. Patients with only a single draw had that value used directly. This approach was selected to capture upper-end inflammatory burden over time while reducing sensitivity to isolated extreme values that may reflect acute illness or measurement variability. Time-weighted approaches were considered but the irregular CBC sampling made them less reliable.

### Definitions of Aortic Growth

Aortic growth was treated as a binary variable for analysis. Growth was defined as an increase of ≥2 mm in mid-AsAo diameter during the surveillance interval, a threshold that was chosen because this degree of change exceeds typical inter-scan measurement variability.^14,15^ Intervals shorter than six months were excluded.

### Statistical Analysis

Associations between inflammatory markers and aortic growth were evaluated at two analytical levels. At the patient level, univariable logistic regression was performed using the 95th percentile of each marker summarized across the entire surveillance window, with one observation per patient. At the interval level, logistic regression was performed using generalized estimating equations (GEE) analyzing each consecutive scan pair independently while accounting for within-patient correlation.

Multivariable logistic regression was then performed to assess the independent association between each marker and binary aortic growth, after adjustment for age, sex, body mass index (BMI), baseline mid-AsAo diameter, systolic blood pressure, diabetes mellitus, and HTAD. Results are reported as odds ratios (OR) with 95% confidence intervals (CI).

To focus on the subgroup of patients with guideline-defined ascending aortic dilation, models were stratified at a baseline mid-AsAo diameter of 40 mm (<40 mm vs ≥40 mm), the threshold at which imaging surveillance is recommended,^1^ and effect modification was tested with a marker-by-diameter-group interaction term. Sensitivity analyses (1) excluded intervals with extreme NLR values (>99th percentile) and (2) substituted the mean, median, and maximum for the 95th percentile of each marker. GEE models used an exchangeable working correlation structure clustered by patient. Continuous variables are reported as median (interquartile range [IQR]) and were compared using the Mann–Whitney U test; categorical variables are reported as n (%) and were compared using the chi-square test. A two-sided p<0.05 was considered statistically significant.

## Results

### Study Population

Of 3,262 patients with serial CT imaging of the native ascending aorta, CBC data during surveillance were available for RDW in 1,882, NLR in 1,471, and SII in 1,468. The population included for analysis consisted of 1,882 patients, of whom 67.3% were male, with a median age of 63.1 years (IQR 53.4–70.9), median baseline mid-AsAo diameter of 40.0 mm (IQR 36.0–43.3), and median follow-up of 4.1 years (IQR 2.0–7.0). Hypertension was present in 90.2% of patients and diabetes in 30.8%. Baseline characteristics are presented in Table 1.

**Table 1.** Baseline characteristics of the study cohort by aortic growth status.

| Variable | Total (N=1882) | Growth (N=192) | No Growth (N=1690) | p-value |
|---|---|---|---|---|
| **Demographics** |  |  |  |  |
| Male, n (%) | 1267 (67.3%) | 128 (66.7%) | 1139 (67.4%) | 0.902 |
| Age (years), median [IQR] | 63.1 [53.4–70.9] | 64.1 [49.1–72.0] | 63.1 [53.8–70.8] | 0.463 |
| Height (cm), median [IQR] | 175.3 [165.1–181.0] | 175.3 [165.1–182.1] | 175.3 [165.9–181.0] | 0.973 |
| Weight (kg), median [IQR] | 88.2 [75.0–102.7] | 86.2 [72.2–106.4] | 88.4 [75.2–102.5] | 0.541 |
| BMI (kg/m²), median [IQR] | 28.9 [25.6–33.2] | 28.6 [25.1–33.1] | 28.9 [25.7–33.2] | 0.4 |
| **Hemodynamics** |  |  |  |  |
| Systolic BP (mmHg), median [IQR] | 128.0 [121.0–135.0] | 128.0 [122.0–135.0] | 128.0 [121.0–135.4] | 0.563 |
| Diastolic BP (mmHg), median [IQR] | 72.0 [67.0–77.0] | 72.0 [67.4–77.6] | 72.0 [67.0–77.0] | 0.346 |
| **Aortic Imaging** |  |  |  |  |
| Baseline Diameter (mm), median [IQR] | 40.0 [36.0–43.3] | 39.0 [35.0–44.0] | 40.0 [36.0–43.2] | 0.195 |
| Number of CT Scans, median [IQR] | 3.0 [2.0–5.0] | 3.0 [2.0–4.2] | 3.0 [2.0–5.0] | 0.849 |
| Follow-up (years), median [IQR] | 4.1 [2.0–7.0] | 3.6 [1.8–5.3] | 4.2 [2.0–7.2] | 0.006 |
| **Comorbidities** |  |  |  |  |
| Diabetes, n (%) | 580 (30.8%) | 52 (27.1%) | 528 (31.2%) | 0.271 |
| Hypertension, n (%) | 1698 (90.2%) | 170 (88.5%) | 1528 (90.4%) | 0.484 |
| Marfan Syndrome, n (%) | 42 (2.2%) | 7 (3.6%) | 35 (2.1%) | 0.253 |
| HTAD, n (%) | 56 (3.0%) | 10 (5.2%) | 46 (2.7%) | 0.09 |
| **Inflammatory Markers Across All Surveillance (95th percentile)** |  |  |  |  |
| RDW (%), median [IQR] | 13.8 [13.1–15.0] | 13.9 [12.9–15.6] | 13.8 [13.1–14.9] | 0.336 |
| NLR, median [IQR] | 3.7 [2.4–7.0] | 4.4 [2.5–7.7] | 3.7 [2.4–6.9] | 0.156 |
| SII, median [IQR] | 762.2 [465.8–1483.2] | 825.9 [524.2–1647.8] | 753.8 [459.6–1463.3] | 0.142 |
| **Interval-level Characteristics** |  |  |  |  |
| Interval Duration (years), median [IQR] | 1.8 [1.0–3.0] | 2.0 [1.0–3.4] | 1.7 [1.0–3.0] | 0.017 |

*Data are median (interquartile range) for continuous variables and n (%) for categorical variables. IQR = interquartile range. Continuous variables compared using Mann–Whitney U test. Categorical variables compared using chi-square test. HTAD = heritable thoracic aortic disease. SII = systemic immune-inflammation index. NLR = neutrophil-to-lymphocyte ratio. RDW = red cell distribution width. BMI = body mass index. BP = blood pressure. CT = computed tomography.*

### Inflammatory Markers and Aortic Growth

During surveillance, 192 patients (10.2%) met criteria for aortic growth, with growth occurring across 287 discrete intervals (10.2%). On patient-level univariable logistic regression, no inflammatory marker was significantly associated with aortic growth (RDW OR=1.056, 95% CI=0.990–1.127, p=0.100; NLR OR=1.014, 95% CI=0.993–1.036, p=0.204; SII OR=1.005, 95% CI=0.997–1.013, p=0.216). At the interval level, GEE logistic regression similarly demonstrated no significant association for any marker (RDW OR=1.046, 95% CI=0.984–1.112, p=0.148; NLR OR=1.016, 95% CI=0.996–1.036, p=0.117; SII OR=1.005, 95% CI=0.998–1.012, p=0.196). In a sensitivity analysis excluding intervals with extreme NLR values (>99th percentile; 22 intervals excluded), the interval-level association between NLR and aortic growth remained non-significant (NLR OR=1.023, 95% CI=0.991–1.056, p=0.160).

On multivariable logistic regression, RDW was independently associated with aortic growth at the patient level after adjustment for mid-AsAo diameter, systolic blood pressure, diabetes mellitus, HTAD, and demographics (OR=1.071, 95% CI=1.002–1.144, p=0.043; Figure 1). At the interval level, this association was confirmed on GEE logistic regression after the same adjustment (OR=1.075, 95% CI=1.012–1.141, p=0.019). NLR and SII were not independently associated with aortic growth on multivariable analysis at either level (NLR p=0.080; SII p=0.164; Figures S1 and S2).

**Figure 1.**
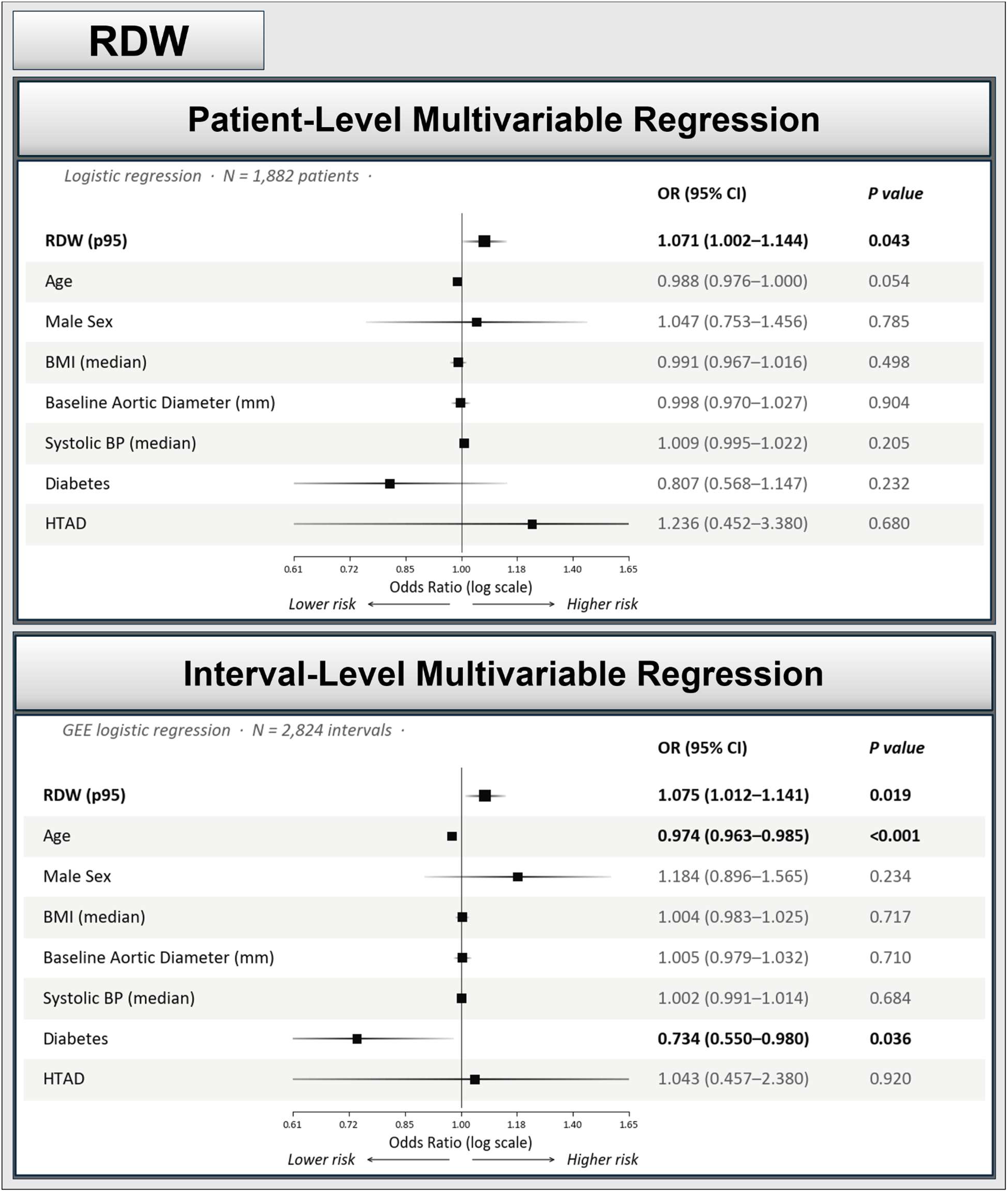
Multivariable Logistic Regression Analyses of RDW and Ascending Aortic Growth at the Patient and Interval Level. Forest plots showing odds ratios (95% CI) for aortic growth from multivariable models with RDW as the primary predictor (top row, bold), adjusted for age, sex, BMI, baseline aortic diameter, systolic blood pressure, diabetes, and HTAD. Top panel: patient-level multivariable logistic regression (N=1,882 patients). Bottom panel: interval-level multivariable GEE logistic regression with exchangeable correlation clustered by patient (N=2,824 intervals). Squares represent point estimates; horizontal lines represent 95% confidence intervals. BMI = body mass index; GEE = generalized estimating equations; HTAD = heritable thoracic aortic disease; RDW = red cell distribution width.

### Ascending Aortic Dilation Subgroup

We next evaluated whether the association between inflammatory markers and aortic growth differed between patients with and without guideline-defined ascending aortic dilation (≥40 mm vs <40 mm) at both the interval and patient levels (Table 2). At the interval level, none of the three markers were associated with growth among aortas <40 mm (RDW OR=1.02, 95% CI=0.94–1.11; NLR OR=1.01, 95% CI=0.99–1.02; SII OR=1.00, 95% CI=0.99–1.01). In contrast, all three markers were associated with growth among aortas ≥40 mm (RDW OR=1.14, 95% CI=1.04–1.24, p=0.004; NLR OR=1.05, 95% CI=1.02–1.07, p<0.001; SII OR=1.01, 95% CI=1.00–1.03, p=0.014). The interaction between baseline diameter and NLR was statistically significant (p=0.01). For RDW and SII, the change in OR point estimates between non-dilated and dilated groups did not reach statistical significance (p=0.10 for both). At the patient level, this pattern was not reproduced. RDW was associated with growth among aortas <40 mm (OR=1.10, 95% CI=1.01–1.19) but not ≥40 mm, and no marker showed evidence of effect modification (all p for interaction >0.45).

**Table 2.** Association Between CBC-Derived Inflammatory Markers and Aortic Growth, Stratified by Baseline Ascending Aortic Diameter (<40 mm vs ≥40 mm).

| Marker | <40 mm OR (95% CI) | ≥40 mm OR (95% CI) | p-value for interaction |
|---|---|---|---|
| **Patient Level** |  |  |  |
| RDW | 1.10 (1.01–1.19) | 1.03 (0.92–1.15) | 0.46 |
| NLR | 1.02 (0.99–1.05) | 1.02 (0.98–1.05) | 0.89 |
| SII (per 100) | 1.01 (0.99–1.01) | 1.01 (1.00–1.03) | 0.6 |
| **Interval level** |  |  |  |
| RDW | 1.02 (0.94–1.11) | 1.14 (1.04–1.24) | 0.1 |
| NLR | 1.01 (0.99–1.02) | 1.05 (1.02–1.07) | 0.01 |
| SII (per 100) | 1.00 (0.99–1.01) | 1.01 (1.00–1.03) | 0.1 |

*Odds ratios (95% CI) are derived from multivariable models adjusted for age, sex, BMI, systolic blood pressure, diabetes, and HTAD and are reported separately by baseline aortic diameter (<40 vs ≥40 mm). Interval-level models use GEE with exchangeable correlation clustered by patient; patient-level models use standard multivariable logistic regression. P-value for interaction tests whether the association between the marker and growth differs between the two diameter groups. SII expressed per 100-unit increase. BMI = body mass index. CI = confidence interval. GEE = generalized estimating equations. HTAD = heritable thoracic aortic disease. NLR = neutrophil-to-lymphocyte ratio. OR = odds ratio. RDW = red cell distribution width. SII = systemic immune-inflammation index.*

### Sensitivity Analyses

Another sensitivity analysis substituting mean, median, and maximum values for the 95th percentile yielded directionally consistent associations for all three markers at both the patient and interval level (Table S1), with the RDW association remaining statistically significant under several alternative summary statistics at the interval level.

## Discussion

In this retrospective analysis of 1,882 patients undergoing serial CT surveillance of the mid-AsAo, comprising 2,824 discrete imaging intervals and 9,371 cumulative patient-years of follow-up, RDW was modestly but independently associated with aortic growth at both the patient level and the interval level. Neither NLR nor SII was associated with growth in the overall cohort at either level of analysis. However, among patients with guideline-defined ascending aortic dilation (≥40 mm), the population under clinical surveillance, all three markers were associated with interval growth, whereas none were among nondilated aortas. This concordance is notable because the markers capture partly distinct biology; RDW reflects inflammation-related disturbance of erythropoiesis accumulated over months, whereas NLR and SII reflect leukocyte and platelet responses. A consistent signal across erythrocyte- and leukocyte-based indices supports an inflammatory contribution to growth once aortic dilation is established. To our knowledge, this is among the largest longitudinal analyses of circulating inflammatory markers and ascending aortic growth to date. A further strength is that, instead of relying on a single value, we related the inflammatory burden within each surveillance interval to the aortic growth observed over that same interval, allowing inflammatory markers to be temporally aligned to the interval in which growth occurred.

Medial degeneration of the ascending aorta is a chronic process. Contractile vascular smooth muscle cells dedifferentiate toward a synthetic phenotype, increase matrix metalloproteinase and cytokine production, and degrade the elastic lamellae over months to years.^5–7^ Consistent with this, murine models of Marfan-associated aneurysm have demonstrated that adventitial inflammation, characterized by progressive immune cell infiltration, tracks closely with aneurysm progression and precedes rupture.^16^ Adaptive immunity may also contribute, as B cells accumulate in the adventitia of dilated human ascending aortas and loss or depletion of B cells attenuates aneurysm development in experimental models, further supporting a mechanistic link between chronic inflammatory activity and aortic wall remodeling.^17–19^ A circulating marker that tracks this process must reflect sustained inflammatory activity. RDW quantifies the variation in circulating red cell volume and rises when erythropoiesis produces a more heterogeneous red cell population, with chronic inflammation as a driver of this heterogeneity. Sustained interleukin-6 signaling raises hepcidin, which sequesters iron within macrophages,^12,20^ while inflammatory cytokines blunt erythropoietin responsiveness and impair erythroid maturation, and the resulting anisocytosis accumulates across the erythrocyte lifespan of roughly 120 days.^11^ RDW therefore behaves as a downstream marker of months of low-grade inflammation. Its independent association with ascending aortic growth supports the premise that chronic inflammatory activity contributes to aortic remodeling and disease progression. The stronger association observed among patients with aortic dilation is consistent with the expectation that chronic inflammatory pathways matter more once structural aortic disease is established and the wall is actively remodeling. The same reasoning explains why RDW carries prognostic weight across heart failure,^10^ stable coronary artery disease,^21^ stroke,^22^ and abdominal aortic aneurysm.^23^

RDW was the only marker of the described pathway available at scale in our data. Markers positioned upstream in this chain may reflect the inflammatory signal more directly. Interleukin-6 (IL-6) is an important mediator of the inflammatory response, while high-sensitivity C-reactive protein (hs-CRP) is a downstream acute-phase marker influenced by IL-6 signaling. Each carries prior support in aortic disease, with a C-reactive protein to IL-6 ratio tracking uncomplicated thoracic aortic aneurysm size^24^ and elevated hs-CRP predicting incident abdominal aortic aneurysm in a prospective population cohort.^25^ We examined these markers in our data. hs-CRP was available during surveillance in 58 patients and IL-6 in 3 patients. These data were too sparse and too selectively obtained to support meaningful inference across a heterogeneous surveillance population. Prospective sampling with a standardized inflammatory panel at fixed surveillance intervals would place these markers on equal footing with imaging and test the chronic-inflammation hypothesis on its own terms.

The null results for NLR and SII in the overall cohort are consistent with the same hypothesis. NLR derives from neutrophil and lymphocyte counts, and SII additionally incorporates platelets, all of which shift within hours in response to infection, trauma, corticosteroids, surgery, and physiologic stress.^26^ A cohort sampled opportunistically across years captures many such episodes, and the variance they introduce reflects transient inflammation remote from medial degeneration. We summarized each marker by its 95th percentile across the interval, which captures peak inflammatory burden while remaining more stable than a single maximum value. This reduces the influence of any one extreme draw, yet the 95th percentile still samples the upper end of each marker’s distribution. For NLR and SII, whose upper values are composed largely of transient elevations, this statistic may capture acute events more than sustained inflammatory tone. In patients with established dilation, in whom medial remodeling is presumably more active, sustained inflammatory activity may be large enough to emerge above this background variability, which may in part explain why NLR and SII were associated with growth only in this group.

NLR and SII have previously been linked to all-cause and cardiovascular mortality in large cohorts,^8,9^ yet neither examined aortic growth. Two prior studies examined CBC-derived indices in thoracic aortic aneurysm. One cross-sectional study reported higher RDW-to-platelet and mean platelet volume ratios in aneurysm patients compared to non-dilated controls.^27^ This is broadly consistent with a link between RDW and ascending aortic disease, though the study addressed a different question. Their study examined whether RDW is elevated in an already established disease, whereas our study examined whether absolute RDW values predict growth over longitudinal follow-up. A second study found that a preoperative NLR above 3.5 was significantly more common among patients with symptomatic or ruptured aTAA, suggesting that NLR may be associated with more advanced or acute aortic disease.^28^ In the overall cohort, we found no association between NLR and aortic growth during longitudinal outpatient surveillance in an asymptomatic population, although NLR was associated with interval growth among patients with aortic dilation. This discrepancy is likely related to differences in patient population and clinical setting. That study measured NLR once, immediately before surgery, when levels may have been influenced by the acute presentation, whereas our study evaluated repeated NLR measurements collected over several years and examined their association with subsequent aortic growth. While NLR may have value as a marker of symptomatic or ruptured disease, our findings do not support its use as a standalone predictor of growth across all patients under surveillance, although it may carry signal in those with established dilation.

### Limitations

Our study is not without limitations. First, our data are retrospective and from a single center, limiting generalizability. Second, the use of electronic medical records may not fully capture the comorbidities, inflammatory conditions, infections, medication changes, and interval procedures that influence the biomarkers in this study. Third, and relatedly, CBC data were not uniformly available, and patients with more laboratory data may have had more severe or chronic disease and greater health care contact. Fourth, growth was modeled as a binary rather than continuous outcome, precluding assessment of a dose-response relationship. This dichotomous definition was chosen deliberately, since clinical aortic diameter measurements carry substantial intrinsic variability, and annualizing modest changes over short surveillance intervals amplifies this error to produce spurious growth rates. Fifth, the dilation-stratified analyses were secondary, formal effect modification reached significance only for NLR, and because SII is derived from the same neutrophil and lymphocyte counts as NLR, these two indices do not provide fully independent evidence. Our findings, including the RDW association and the signal among dilated aortas, should therefore be considered hypothesis-generating and require validation in an independent, prospectively sampled cohort before clinical application.

## Conclusions

In conclusion, among a large cohort of patients undergoing longitudinal CT surveillance of the ascending aorta, elevated RDW was modestly but independently associated with aortic growth, whereas NLR and SII showed no independent association in the overall cohort. Among patients with guideline-defined ascending aortic dilation, however, all three inflammatory markers were associated with interval growth, a concordant signal that supports a contribution of systemic inflammation to disease progression in the population undergoing surveillance. While the modest effect sizes limit the value of these markers for standalone risk stratification, these associations provide proof of concept that systemic inflammatory activity may contribute to ascending aortic remodeling and support prospective evaluation of more specific markers such as hs-CRP, erythrocyte sedimentation rate, and IL-6, particularly in patients with aortic dilation.

## Acknowledgments

We would like to acknowledge the Cardiovascular Health Improvement Project (CHIP) and MI-AORTA programs at the Frankel Cardiovascular Center for their support of this project through programmatic and database resources.

## Sources of Funding

NSB – David A. Bluemke Professorship, NIH/NHLBI (R01HL170059).

## Disclosures

Dr Burris is entitled to royalties related to licensure of intellectual property and is the founder of and an equity holder in VeriVasc, Inc., which develops aortic imaging software; VeriVasc had no role in this study. Dr Karnam is entitled to royalties related to licensure of intellectual property. The remaining authors have no disclosures to report. The remaining authors have no disclosures to report.

## Supplemental Material

Table S1 and Figures S1–S2.

## Nonstandard Abbreviations and Acronyms

aTAA: Ascending thoracic aortic aneurysm
CBC: Complete blood count
EMR: Electronic medical record
GEE: Generalized estimating equations
HTAD: Heritable thoracic aortic disease
hs-CRP: High-sensitivity C-reactive protein
mid-AsAo: Mid-ascending aorta
NLR: Neutrophil-to-lymphocyte ratio
RDW: Red cell distribution width
SII: Systemic immune-inflammation index

## Notes

### Author Declarations

This retrospective cohort study received approval from the University of Michigan (IRBMED) Institutional Review Board (HUM00133798)

